# Efficacy of Anakinra in the Management of Patients with COVID-19 Infection: A Randomized Clinical Trial

**DOI:** 10.1101/2022.07.04.22277207

**Authors:** Eman Zeyad I. Elmekaty, Aya Maklad, Rawan Abouelhassan, Waqar Munir, Mohamed Izham Mohamed Ibrahim, Arun Nair, Rim Alibrahim, Fatima Iqbal, Ahmad Al Bishawi, Alaaeldin Abdelmajid, Mohamed Aboukamar, Mohammed Abu Khattab, Hussam Al Soub, Muna Al Maslamani

## Abstract

**Background:** The global pandemic of COVID-19 infections continues to grow worldwide, with rising number of deaths day by day. The hyperinflammation state contributes to the multiorgan failure associated with COVID-19 infections. This study aims to explore the efficacy and safety of anakinra in COVID-19 patients with both respiratory distress and cytokine release syndromes.

**Methods:** This was an open-label, multicenter, randomized clinical trial. Patients were randomized in 1:1 ratio to receive standard of care (SOC) alone, or anakinra plus SOC. Adults with confirmed COVID-19 infection with evidence of both respiratory distress and cytokine release syndrome were included. The primary outcome was treatment success at day 14, defined as WHO clinical progression score of ≤3. The primary analysis was based on intention-to-treat population, with p-value of <0.05.

**Results:** A total of 80 patients were enrolled in the study. The mean age was 49.9 years (SD=11.7), with 82.5% (n=66) male patients. The primary outcome was not statistically different (87.5% (n=35) in anakinra group vs. 92.5% (n=37) in SOC group, p=0.712). The majority of reported adverse events were mild in severity and not related to the study treatment. Increased aspartate aminotransferase was the only significant adverse event (35% (n=14) in anakinra group vs. 15% (n=6) in SOC group, p=0.039); yet, was not associated with treatment discontinuation.

**Conclusion:** In patients with severe COVID-19 infection, the addition of anakinra to SOC treatment was not associated with significant improvement in the WHO clinical progression scale. Further studies investigating patients’ subgroups that might benefit from anakinra are warranted. The trial was registered at ClinicalTrials.gov (NCT04643678).

## Introduction

Since its declaration by the World Health Organization (WHO) in March 2020 as a global pandemic, the severe acute respiratory syndrome coronavirus (SARS-CoV-2), also known as COVID-19, continues to spread worldwide with increasing daily fatalities ^[1]^. Subsequently, as of September 2021, the total number of confirmed COVID-19 infections has exceeded 200,000,000 cases, with a total of 4,500,000 recorded deaths globally ^[2]^. Although the majority of confirmed infections present with mild to moderate clinical presentations ranging from the absence of symptoms to the loss of taste or smell, headache, fatigue or fever, COVID-19 may still result in consequent severe sequelae, including acute respiratory distress syndrome (ARDS) and organ dysfunction syndrome along with an estimated mortality rate of 3.4% ^[3,4]^.

The infection acts as an essential source for activating macrophages, resulting in the release of pro-inflammatory cytokines and chemokine macrophages primarily in the lungs, followed by subsequent systemic release. This process is thought to play a major role in the pathophysiology of organ dysfunction syndrome ^[5]^. Furthermore, cytokine storm syndrome, a state of hyper-inflammation that contributes to multiorgan failure, has proven to share considerable biochemical overlap with the hyper-inflammation observed in patients with confirmed COVID-19 infections. Accordingly, recent evidence hypothesized that screening COVID-19 patients for hyper-inflammation and consequently providing management with immunosuppressive medications could improve mortality ^[6,7]^.

Anakinra, a recombinant non-glycosylated form of the human interleukin (IL)-1 receptor antagonist, has demonstrated significant survival benefits in patients with macrophage activation syndrome (MAS) as compared to placebo, with a 28-day mortality rate of 35% and 65%, respectively (p = 0.0006) in a subgroup analysis of patients with hyper-inflammation as a result of severe sepsis ^[8]^. Furthermore, numerous studies revealed that the administration of anakinra was associated with an acceptable safety profile where recent literature reported mild elevations in liver aminotransferases as the most common adverse effect; however, such elevations were not clinically significant ^[9,10]^.

Meanwhile, ongoing trials are being conducted to investigate further the efficacy and safety profile of anakinra in COVID-19 patients exhibiting features of cytokine storm during their illness. Thus far, a variety of published data has shown promising results. However, such findings require extensive assessments in a well-conducted randomized controlled trial to confirm the reported outcomes ^[11,12]^. Currently, there are no approved treatments directly targeted towards ceasing the cytokine storm and its consequences. Thus, this randomized clinical, open-label trial aims to explore the efficacy and safety of anakinra in addition to standard of care (SOC) compared to SOC alone in patients with confirmed COVID-19 infection with respiratory distress and evidence of cytokine release syndrome.

## Methods

### Study design and setting

This study was a prospective randomized clinical, open-labeled, parallel assignments, multicenter trial. Patients were recruited from three clinical sites in Qatar: Communicable Disease Center (CDC), Hazm Mebaireek General Hospital (HMGH), and The Cuban Hospital (TCH) between 30 October 2020 till 30 April 2021. Eligible patients were randomized to receive either anakinra + SOC or SOC alone. The trial protocol and the statistical analysis plan is available online. The trial was approved by the Institutional Review Board at Hamad Medical Corporation (HMC) Medical Research Center (MRC-01-20-1095) and was registered at ClinicalTrials.gov (NCT04643678).

### Participants

Hospitalized adults (age ≥ 18yrs) were eligible for inclusion in the trial if they met the following eligibility criteria: confirmed COVID-19 diagnosis by positive SARS-CoV2 Polymerase Chain Reaction (PCR) test and associated presence of respiratory distress [defined as: PaO2/FiO2 ≤ 300 mm Hg or respiratory Rate (RR) ≥24 breaths/min or SpO2 ≤ 94% at room air], signs of cytokine release syndrome [defined as any of the following at baseline: Ferritin >600 mcg/L at presentation or >300 mcg/l with doubling within 24 hours, LDH >250 IU/L, D-dimers > 1 mg/L, CRP > 70mg/L and rising since last 24h with the absence of bacterial infection, Interleukin-6 level > 10 x UNL (reference range ≤7 pg/ml)], radiological evidence of pneumonia based on chest X-ray and/or computed tomography (CT) scan findings, and a signed informed consent provided by the patient, or by the patient’s legal representative. The exclusion criteria included known serious allergic reactions including anaphylaxis to the study medication or any component of the product, active infectious diseases such as active bacterial infections (defined as the isolation of bacteria from a sterile body site or other body sites and is causing clinical symptoms and signs and therefore are treated with antibiotics), invasive fungal infections, Human Immunodeficiency Virus (HIV), Hepatitis B Virus (HBV) infection, Hepatitis C Virus (HCV) infection, active tuberculosis, patients on immunosuppressants or immunomodulatory drugs or had received any in the past 30 days, neutrophil count below 500 cells/microliter, platelets below 50,000/microliter, pregnant or breastfeeding females. A full list of eligibility criteria is provided in the supplementary materials.

### Randomization and masking

All admissions with a laboratory-confirmed SARS-CoV-2 infection and on respiratory distress were screened for eligibility by the research team. Eligible patients were randomly assigned using simple randomization at an allocation ratio of 1:1 to receive either anakinra plus SOC therapy or SOC therapy alone. A random sequence of numbers was generated by an independent biostatistician using a computerized system. Only the principle investigator was responsible to allocate the participant serial number and inform the research team of the treatment allocation. Each participant was assigned an identification code corresponding to a study treatment group. The study was not blinded due to practical reasons and the differences in the dosage regimens and administrations between the study groups.

### Interventions and Procedures

Patients in the intervention group received anakinra (KINERET^®^, 100mg/0.67 ml in pre-filled syringe) 100 mg subcutaneous (SC) injection every 12 hours for 3 days, then 100 mg SC once daily from day 4 to day 7. The total duration of anakinra was 7 days with a total of 10 doses. Patients with severe renal impairment (creatinine clearance <30 ml/min) had a 50% decrease in dose (dose administered every other day). Patients in the intervention group received anakinra in addition to standard of care therapy except for tocilizumab, due to the contraindication of administering both agents concomitantly. Patients in the comparator group received SOC therapy as per the local treatment guideline as approved by the Ministry of Public Health (MoPH) in Qatar at that time (see supplementary material). According to the treating physician’s judgment, therapy was discontinued, or doses were adjusted based on the safety concerns or issues related to concomitant drug interactions. Apart from the study interventions and related laboratory and diagnostic assessments, patients were treated as part of standard care, with no co-medications, medical procedures, or diet restrictions. The study had pragmatic and adaptive nature, which mimics the usual clinical practice in a real-world setting and allows for changes in the management of COVID-19 patients and SOC therapy, as approved by the MoPH in the country.

After enrollment, a detailed review of the participant’s medical profile was conducted by the research team. Demographic, clinical, radiological, laboratory data were collected from each patient’s profile. The WHO Clinical Progression Scale^[13]^ was used to assess the patient’s clinical condition on a daily basis and was documented in the electronic data collection sheet. The worst score on each particular day was selected and recorded. The data collection sheet was validated independently by two investigators. The list of study procedures, timings, and follow-up is available in the supplementary materials.

All adverse events (AEs) that occurred during the hospital stay and up to 28 days from randomization were reported. Participants were contacted by telephone using standardized telephone script on days 14 and 28 to ascertain information related to AEs. All AE were assessed for severity and graded using the Common Terminology Criteria for Adverse Events (CTCAE) version 5^(14)^. A blinded assessor evaluated and validated the AE’s severity, seriousness, relatedness, and attribution using appropriate scales and/or definitions as per the HMC IRB Standard Operating Procedure (SOP) guidelines.

### Outcomes

The primary outcome was treatment success on day 14, defined as a WHO Clinical Progression score of ≤3 [Ambulatory mild disease: symptomatic, assistance needed]. Secondary outcomes included the duration of mechanical ventilation in ventilated patients up to 14 days, changes in the WHO Clinical Progression Score between day 1 and day 7, viral burden as measured by the change in SARS-CoV-2 PCR Cycle Threshold (CT) at day 7 and day 10-14, time to ICU admission up to 28 days, the incidence of adverse events up to 28 days, length of hospital stay up to 28 days, and all-cause mortality rate at hospital discharge or at 28 days, whichever occurred first.

### Statistical analysis

No published randomized trials evaluated the difference between the two study interventions at the time of study planning to appropriately estimate the mean difference and compute the sample size calculation. Therefore, a sample size of 80 patients was estimated for this RCT with parallel group design based on the hypothesized effect size from published observational studies with 80% power to detect a difference in the primary endpoint of 30%, with a two-sided type I error rate of 0.05 (alpha). Categorical data were expressed by frequency (percentage), while continuous values were expressed as mean ± SD or median and interquartile range (IQR), as appropriate. Data were tested for their normality. Kolmogorov-Smirnov test was carried to test for data normality. Continuous data were examined with the Mann–Whitney U or independent t-test and categorical data with the chi-square and Fisher’s Exact tests. Two-way repeated measure ANOVA analyzed the changes of data over time in the two treatment groups. Factors significant at univariate analysis (p<0.10) were further assessed by the multivariable binary regression analysis. All statistical analyses were carried out using the statistical package SPSS, version 27 (IBM Corp. Released 2020. IBM SPSS Statistics for Windows, Version 27.0. Armonk, NY: IBM Corp). The primary analysis was based on an intention-to-treat population, including all participants randomized.

### Ethics Statement

The trial was approved by the Institutional Review Board at Hamad Medical Corporation (HMC) Medical Research Center (MRC-01-20-1095) and was registered at ClinicalTrials.gov (NCT04643678). The trial was conducted in accordance with Good Clinical Practice guidelines and the principles of the Declaration of Helsinki. An independent institutional review board and ethics committee approved the trial protocol and any subsequent amendments. The safety of the participants and the evaluation of the benefit–risk balance was overseen by an independent data safety monitoring committee. Written informed consent was obtained from all participants or legally authorized representatives before initiating any trial-related procedures. No additional administrative permissions were required to access the raw data. All data used in this study were fully anonymized before their use.

## Results

### Participants

Between 30 October 2020 and 28 February 2021, 327 patients were screened for eligibility and 80 patients were enrolled and randomized to the two study groups: 40 patients in anakinra group and 40 patients in the SOC group. One patient in the SOC group was lost to follow-up and discharged from the hospital for social reasons [Figure 1]. All patients randomized were included in the intention to treat analysis. As enrolled patients were hospitalized during the treatment course, the compliance with study medication was 100%.

**Figure 1:**
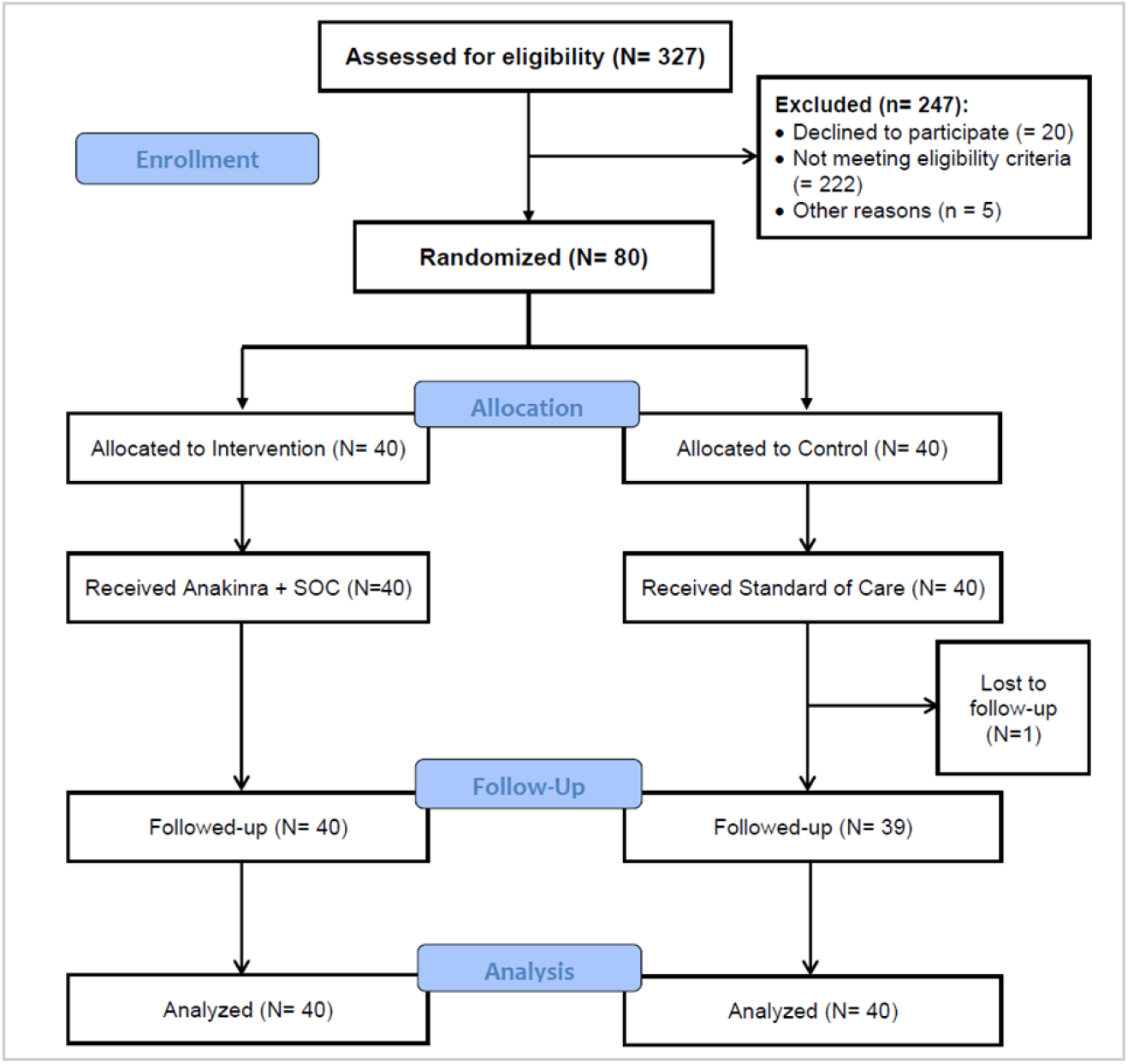
Flow diagram

The baseline characteristics of the patients were summarized in table 1. No significant differences were found (p>0.05) between the two treatment groups. The complete list of participants demographics, clinical data, and concomitant medications is available in the supplementary materials.

**Table 1:**
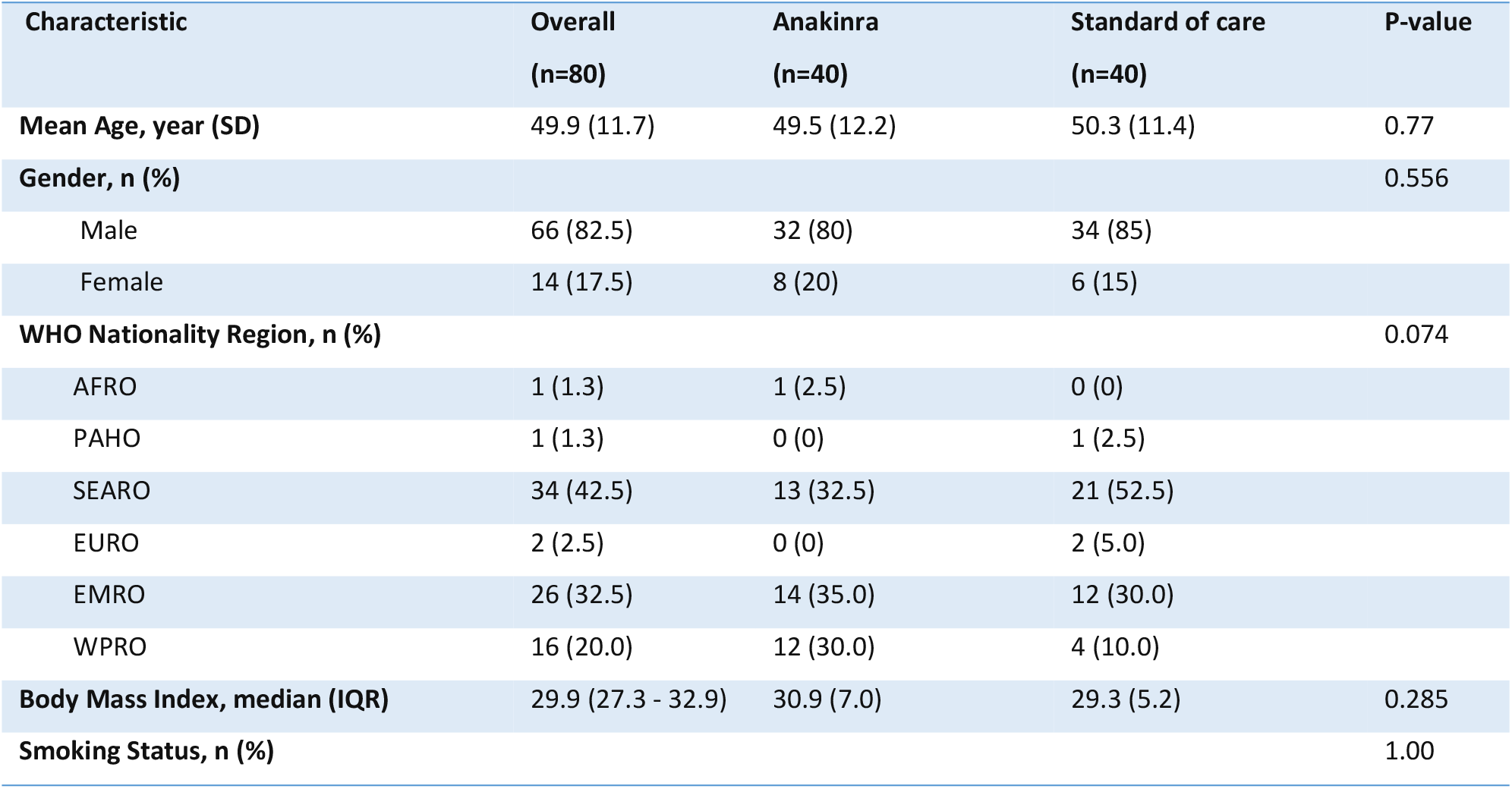
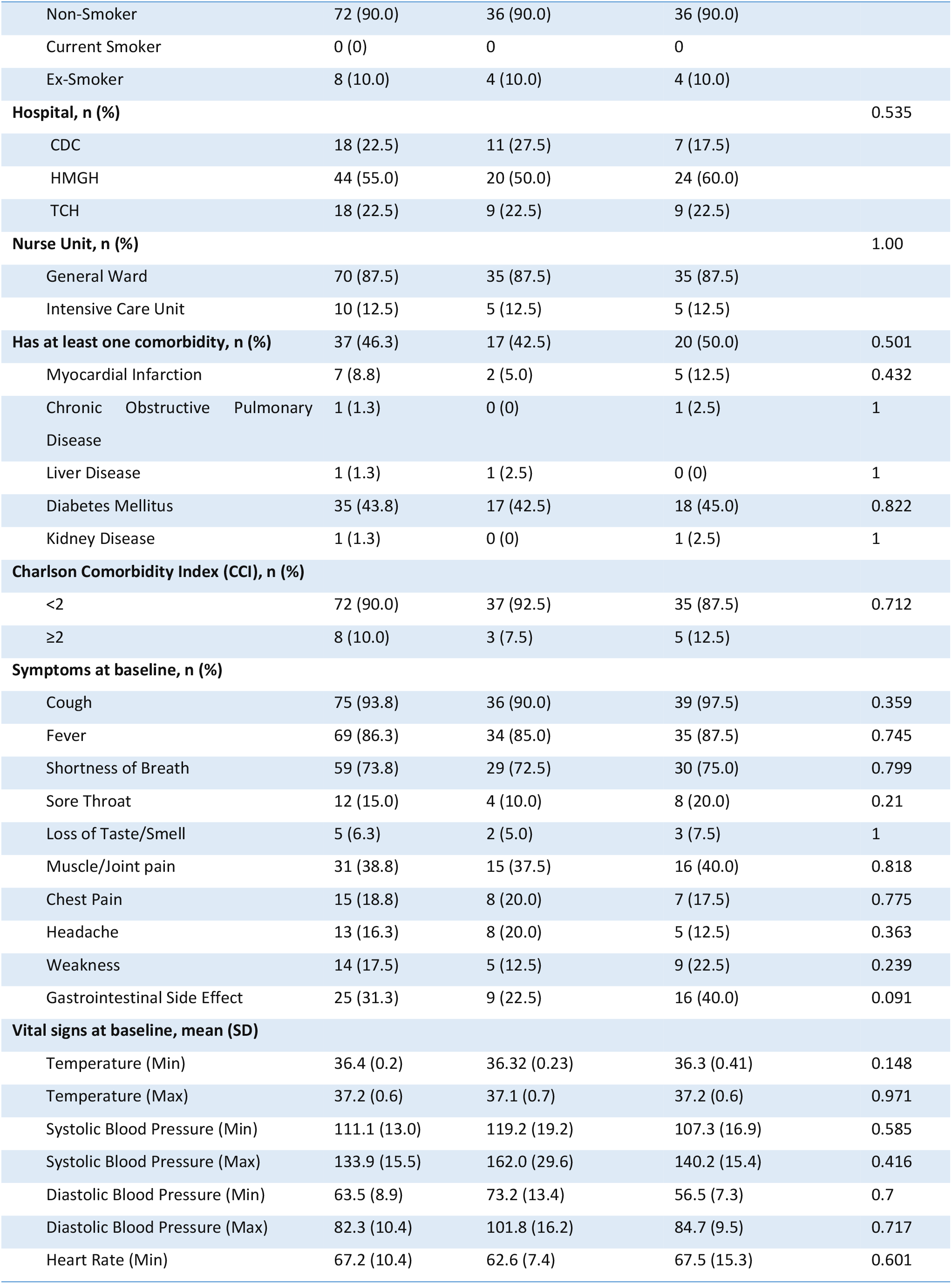
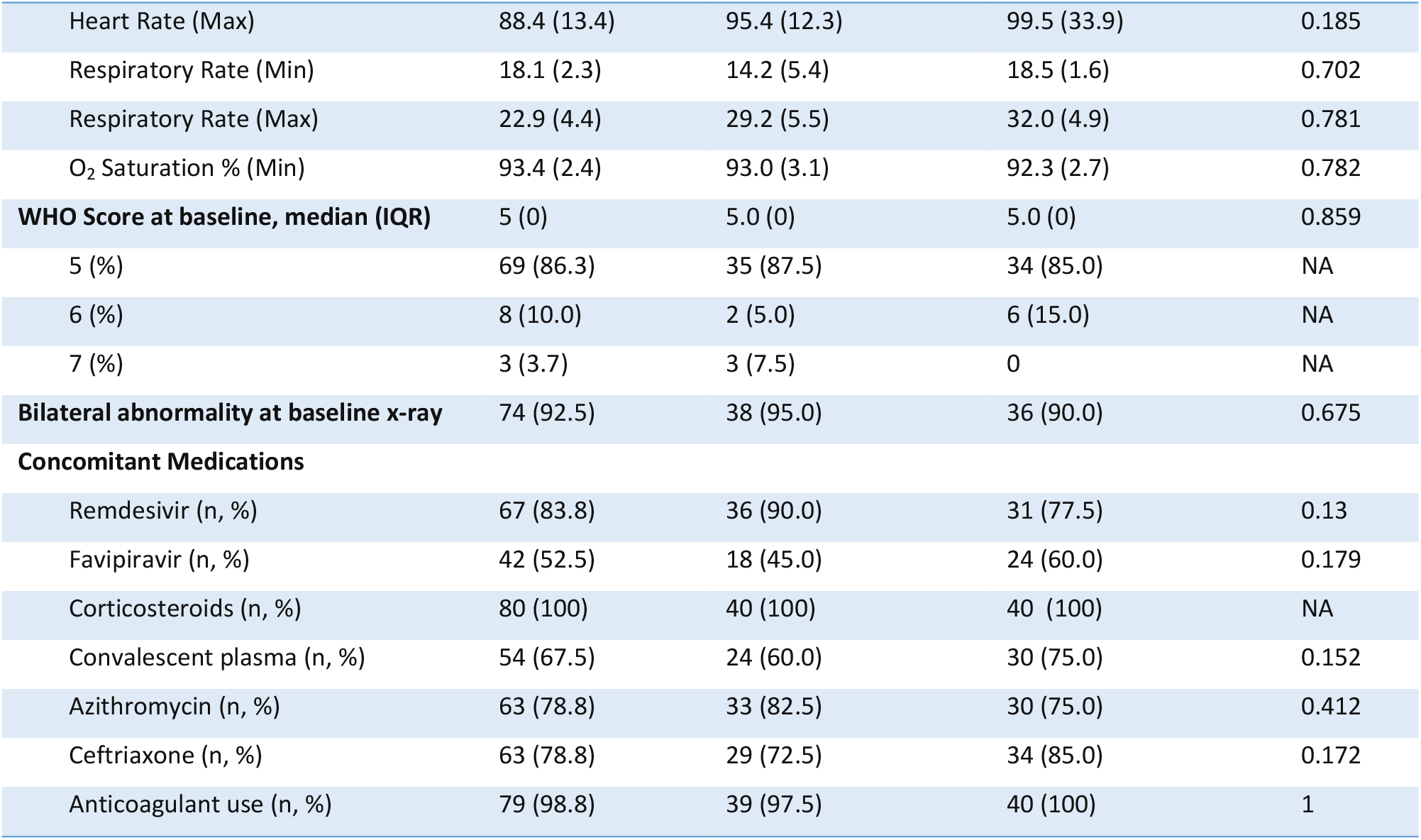
Baseline characteristics of the study population.

The change in vital signs and laboratory data over time (i.e. from baseline to day 14) were illustrated in figures 1-36 in the supplementary materials. No significant difference over time in the two treatment groups was noted except for the lymphocyte count (marginal; p=0.046). Figure.2 demonstrates a significant change in WHO score (p=0.000) over 14 days.

**Figure.2:**
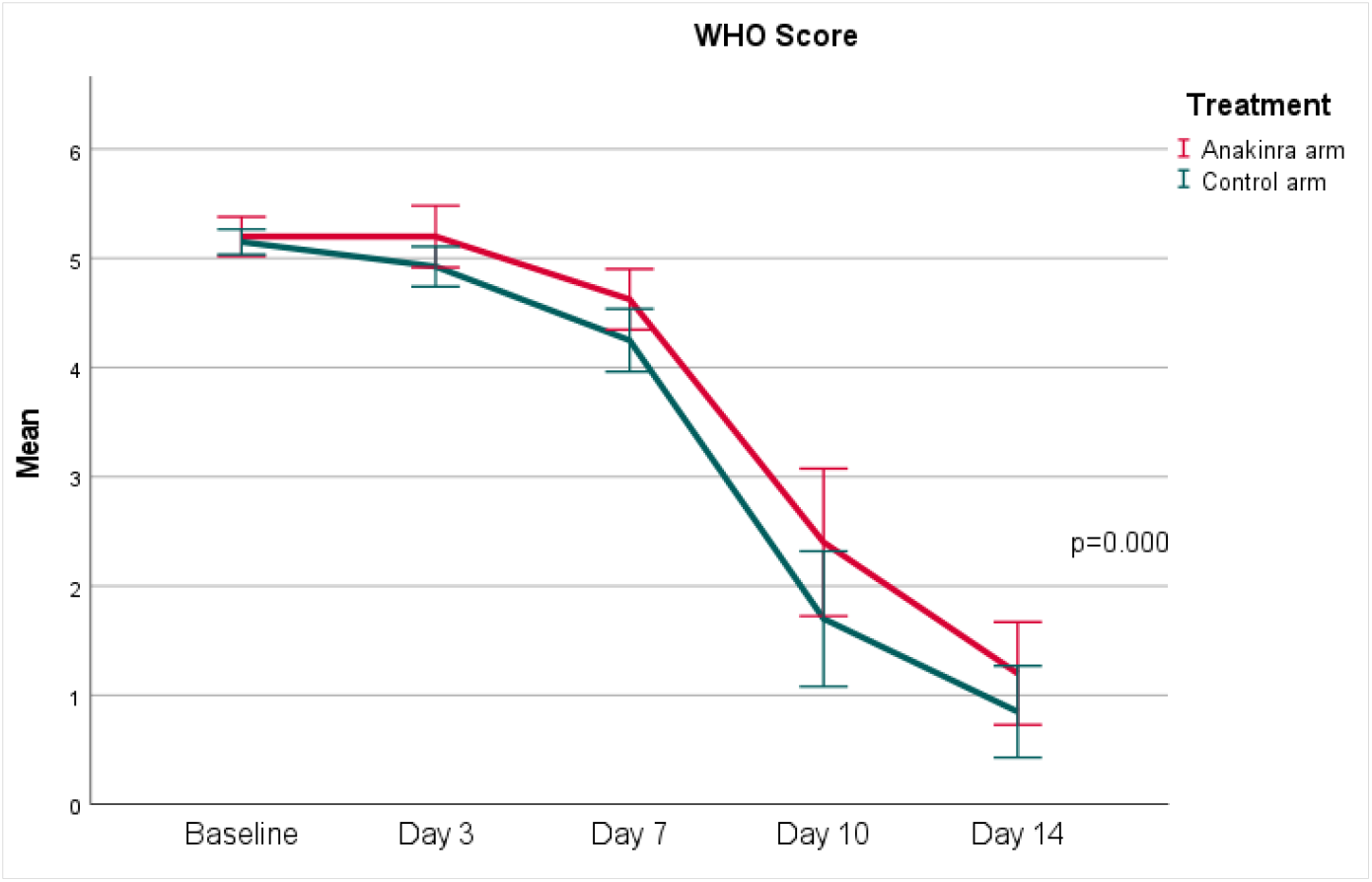
Change in the WHO Clinical progression Score between days 1 and day 14

### Outcomes

Table 2 below illustrates the results of the primary and secondary outcomes for the two treatment groups. The percentage of patients with treatment success at day 14 was not significantly different between the two groups (p=0.712). None of the secondary outcomes were found to be statistically significant.

**Table 2:** Results of the primary and secondary outcomes.

| Outcome | Overall<br>(n=80) | Anakinra<br>(n=40) | Standard of care<br>(n=40) | P-value |
| --- | --- | --- | --- | --- |
| **Primary outcome:** |  |  |  |  |
| Percentage of patients with treatment success at day 14, n (%) | 72 (90.0) | 35 (87.5) | 37 (92.5) | 0.712 |
| **Secondary outcomes:** |  |  |  |  |
| Duration of mechanical ventilation in ventilated patients up to day 14, median days (IQR) | 6.50 (2.0) | 6.50 (2.0) | NA | NA |
| Change in WHO score from baseline to day 7, median (IQR) | 1 (0 - 1) | 1 (1) | 1 (1) | 0.211 |
| Change in viral burden (CT value) from baseline to day 7, median (IQR) | 13.1 (7.8 - 23.2) | 6.1 (14.9) | 9.8 (7.7) | 0.09 |
| Change in viral burden (CT value) from baseline to day 10-14, median (IQR) | 17.6 (8.5 - 23.9) | 15.8 (16.0) | 17.6 (17.0) | 0.923 |
| Time to ICU admission in non-ICU patients up to 28 days, median time (IQR) | 6.0 (4.0 – 15.0) | 7.5 (13.0) | 6.0 (15.3) | 1.00 |
| Length of hospital Stay, median days (IQR) | 10.0 (8.0 - 13.0) | 10.0 (4) | 10.0 (4) | 0.064 |
| All-cause mortality, n (%) | 1 (1.25) | 0 (0) | 1 (2.5) | 1.000 |

The association between baseline characteristics and the primary outcome was not statistically significant between the two groups (refer to supplementary materials).

### Safety Outcomes

Two DSMB meetings were conducted as planned, in which the study’s progress, execution, results, and adverse event reports were reviewed by the committee members. At each meeting, the decision was made to proceed with the study accrual. A total of 377 adverse events were identified in the study. All patients had at least one adverse event, with an average of 4.7 events per patient. The majority of adverse events (61%) were mild in severity and did not cause harm to patients. Furthermore, 78% of the reported adverse events were judged by a blinded assessor to be not related to the study, 21% “possibly related”, and 1% “definitely related” to the study treatment. The most common adverse events were hypertension (65%), followed by increase in alanine aminotransferase, hypoalbuminemia, and sinus bradycardia (43.8% for the three events). Only increased aspartate aminotransferase level was found to be significantly higher in the anakinra group compared to the SOC group (35% vs. 15%), respectively, (p=0.039). Four serious adverse events were reported in the study, two in the anakinra group and two in the SOC group. To elaborate, one patient in the SOC died 23 days after randomization due to worsening of his pre-existing medical condition, while the second patient had a prolonged hospital stay due to diarrhea, in which both were judged to be unrelated to study interventions. In the anakinra group, both patients were re-admitted to the hospital for 3-6 days after two weeks of randomization due to suspected respiratory tract infections, which were judged to be possibly related to the study interventions. Table 3 summarizes the common adverse events reported in the study.

**Table 3:** Reported Adverse Events for Study Groups.

| Adverse events, n (%) | Overall<br>(n=80) | Anakinra<br>(n=40) | Standard of care<br>(n=40) | P-value |
| --- | --- | --- | --- | --- |
| **Incidence of at least one AE** | 80 (100) | 40 (50) | 40 (50) | NA |
| **Average number of AE per patient** | 4.7 | 4.7 | 4.72 | 0.034 |
| **Serious AE** | 4 (5) | 2 (2.5) | 2 (2.5) | NA |
| **Number of Adverse events** | 377/377 (100) | 201/377 (53) | 176/377 (47) | NA |
| Severe AE (grade 4-5) | 1/377 (0) | 0/377 (0) | 1/377 (0) | NA |
| Mild-Moderate AE (grade 1-3) | 376/ 377 (100) | 201/377 (53) | 175/377 (46) | 0.034 |
| **Relatedness to study intervention** |  |  |  |  |
| Not related | 294/377 (78) | 154/377 (41) | 140/377 (37) | 0.863 |
| Possibility related | 79/377 (21) | 44/377 (12) | 35/377 (9) | 0.534 |
| Probably related | 2/377 (1) | 1/377 (0) | 1/377 (0) | NA |
| Definitely related | 2/377 (1) | 2/377 (1) | 0/377 (0) | NA |
| **Common AE (incidence > 5%)** |  |  |  |  |
| Hypertension | 52 (65.0) | 27 (67.5) | 25 (62.5) | 0.639 |
| Alanine aminotransferase increased | 35 (43.8) | 19 (47.5) | 16 (40.0) | 0.499 |
| Hypoalbuminemia | 35 (43.8) | 18 (45.0) | 17 (42.5) | 0.822 |
| Sinus Bradycardia | 35 (43.8) | 16 (40.0) | 19 (47.5) | 0.499 |
| Hypotension | 32 (40.0) | 19 (47.5) | 13 (32.5) | 0.171 |
| Anemia | 21 (26.3) | 8 (20.0) | 13 (32.5) | 0.204 |
| Aspartate aminotransferase increased | 20 (25.0) | 14 (35.0) | 6 (15.0) | 0.039 |
| Hyponatremia | 20 (25.0) | 11 (27.5) | 9 (22.5) | 0.606 |
| Hyperglycemia | 17 (21.3) | 9 (22.5) | 8 (20.0) | 1 |
| Lymphocyte count decreased | 15 (18.8) | 8 (20.0) | 7 (17.5) | 0.775 |
| ECG QT corrected interval prolonged | 9 (11.3) | 5 (12.5) | 4 (10.0) | 1 |
| Alkaline phosphatase increased | 8 (10.0) | 5 (12.5) | 3 (7.5) | 0.712 |
| Diarrhea | 8 (10.0) | 3 (7.5) | 5 (12.5) | 0.712 |
| Hyperkalemia | 7 (8.8) | 5 (12.5) | 2 (5.0) | 0.432 |
| Hypokalemia | 7 (8.8) | 3 (7.5) | 4 (10.0) | 1 |
| Sinus tachycardia | 6 (7.5) | 4 (10.0) | 2 (5.0) | 0.675 |
| Lymphocyte count increased | 4 (5.2) | 2 (5.0) | 2 (5.0) | 1.000 |
| Creatinine increased | 4 (5.2) | 2 (5.0) | 2 (5.0) | 1.000 |

## Discussion

This open-label randomized multicenter clinical trial conducted in Qatar between October 2020 and April 2021, found that anakinra administered SC was not more efficacious than the SOC therapy in patients with COVID-19 pneumonia in terms of achieving higher rates of treatment success; defined as a WHO clinical progression score of 3 or less. The use of anakinra was also not associated with a significant change in the WHO clinical progression score or reducing viral burden at 14 days when compared to usual care. The length of stay and the time to ICU admission for non-ICU patients did not differ between the two groups. The overall mortality rate was low in this trial (1.25%) as only one death was reported in the SOC group.

The main findings in our trial are consistent with that of the randomized clinical trial by the CORIMUNO collaborative group from France ^[15]^, which is the only published RCT investigating the use of anakinra to this date. The CORIMUNO-ANA-1 trial was an open-label, multicenter, randomized controlled trial of anakinra administered intravenously (IV) for 5 days compared to usual care to assess its survival benefits and the need for mechanical ventilation at day 4. Similar to our population, the CORIMUNO trial included patients with a WHO clinical progression score of ≥ 5. The two primary outcomes were not significant, and the need for mechanical or non-invasive ventilation at day 14 was also not different in both groups. Overall mortality at days 28 and 90 did not differ between the groups, and as a result the trial was prematurely stopped for futility.

Anakinra is FDA approved for use in rheumatoid arthritis, neonatal-onset multisystem inflammatory disease, and deficiency of IL-1 receptor antagonist. It has also been utilized in various other clinical situations, most notably its off-label use in the treatment of hemophagocytic lympho-histiocytosis and cytokine storms ^[16,17]^. The hyperinflammation seen in patients with COVID-19 pneumonia is associated with severe disease presentation and is often identifiable by elevation of common inflammatory markers such CRP, IL-6, and procalcitonin, as well as other cytokines ^[18]^. In addition, earlier studies suggested a significant role of raised cytokines, including IL-1, in severe COVID-19 pneumonia, which hypnotized a potential role of IL-1 inhibitors in patients with COVID-19 pneumonia ^[6,17]^.

Several case series and observational studies suggested the potential clinical improvement and mortality benefit of anakinra use in COVID-19 pneumonia ^[19-21]^. A recent meta-analysis of 15 observational and interventional studies with a total of 3530 patients suggested that there is an overall mortality benefit with the use of anakinra when compared to usual care ^[22]^. Evidence also suggested that anakinra can prevent the progression to severe disease and reduce the need for mechanical ventilation. However, these studies were limited by the lack of randomization and the potential for selection and confounding bias, as well as the significant heterogeneity observed in the meta-analysis outcomes.

Unfortunately, the mortality benefit observed in the previous observational studies was not observed in our trial or the CORIMUNO-ANA-1 trial ^[15]^. Despite including patients with severe COVID-19 pneumonia, this trial reported only one patient death (1.25%) in the SOC group. On the other hand, the CORIMUNO-ANA-1 trial reported an overall mortality rate of 28 % in each group. The advanced SOC may partly explain the low mortality rate in this trial compared to the CORIMUNO-ANA-1 trial. To explain, in the three sites, SOC for all patients included administration of dexamethasone and remdesivir as per the national treatment guidelines. However, none of the CORIMUNO-ANA-1 trial patients received antivirals like remdesivir or favipravir, and dexamethasone was not established as part of routine care for moderate to severe pneumonia by then. Dexamethasone in the RECOVERY trial showed mortality benefit and, eventually, became standard of care globally for treating severe COVID-19^[23]^. The Infectious Disease Society of America treatment guidelines recommend using remdesivir in patients requiring supplemental oxygen and in patients with moderate to severe COVID-19 pneumonia, given its proven clinical benefits in this population ^[24]^.

There is an increased interest to prevent severe respiratory failure and the need for intubation and mechanical ventilation in patients presenting with COVID-19 pneumonia. Small case series from the United States initially showed that IV administration of anakinra prevented the development of severe acute hypoxic respiratory failure^[19]^. Another meta-analysis of seven studies (including the CORIMUNO ANA-1 trial), with a total of 740 patients receiving anakinra, showed that anakinra reduced the need for invasive mechanical ventilation^[22]^. The considerable heterogeneity in these studies as well as the predominantly retrospective and observational nature of those studies, warranted the confirmation of these findings in a randomized clinical trial. However, no significant benefit was observed by either this trial or the CORIMUNO-ANA-1 trial with regards to the rates of invasive mechanical ventilation.

The preferred core outcomes for clinical studies as defined by the WHO working group^[13]^ were used in this trial. Cycling Threshold (CT) is used as a marker of viral load, as it is inversely related to the viral load and provides an indirect method of quantifying the copy number of viral RNA^[25]^. Anakinra did not affect the viral burden when compared to the SOC group in this study. Furthermore, the WHO clinical progression scale is a 10-point ordinal scale from 0 (no disease) to 10 (dead). It was chosen as an objective outcome measure to reduce the variability and heterogeneity seen in the previous studies and allow comparisons with future studies. No significant changes were reported in the baseline of the WHO clinical progression score or the seven-days score between the two groups.

Overall, a higher prevalence of adverse events was reported in this trial as compared to other studies, with every patient experiencing at least one adverse event. The most commonly reported adverse events were hypertension, raised liver transaminases, and hypoalbuminemia. Only AST elevation was significantly higher in the anakinra group. Although AST elevation is one of the possible adverse effects of anakinra, the possibility that the reported increase in this trial might be due to the multiple concomitant medications taken by the patients cannot be denied. Overall, the incidence of severe adverse events did not exceed 5%. The high prevalence of adverse events in our trial may partially be attributed to the robust data collection and the additional concomitant use of antivirals as part of SOC therapy compared to SOC in other studies. Convalescent plasma has also been part of Qatar’s national guidelines, and patients received it as per protocol. The decision was made by the treating physician in line with the criteria set for use as per the national CDC guidelines (refer to supplementary materials).

There is increased awareness of the concept of the possible narrow therapeutic window to treat cytokine storms with cytokine inhibitors as well as the right time to initiate IL-1 inhibitors^[26]^. There is a need to define if certain inflammatory markers or other biomarkers can guide the right timing for the use of anakinra. Soluble urokinase plasminogen activator receptor (SuPAR), a molecular measurable marker, is associated with increased risk of severe respiratory failure and has also been found to be elevated in patients with COVID-19^[27,28]^. An open-label non-randomized interventional trial (SAVE) was done to assess the use of anakinra to prevent progression to severe COVID-19 disease ^[29]^. The trial included 130 patients in the anakinra group and 130 propensity-matched SOC comparator cohort. Patients who had a level higher than 6 microg/l were deemed at risk of developing severe respiratory failure and, thus, were treated with anakinra. Anakinra successfully prevented the need for mechanical ventilation and demonstrated mortality benefits. SAVE trial, although inadequate randomization, provided additional insights that patients with biomarker-guided therapy may be better candidates to receive the specific interleukin inhibitors. This is being investigated by the ongoing placebo-controlled randomized (SAVE-MORE) phase III trial assessing the effects of biomarker-guided use of anakinra in patients. (www.clinicaltrials.gov, NCT04680949)^[27]^. Similarly, another non-randomized study, where ferritin was used as a biomarker to guide therapy, showed significantly improved clinical outcomes, including reduced mortality rates and progression to severe COVID-19 by early administration of therapy either with anakinra alone or in combination with steroids^[30]^.

The route of administration of anakinra used in our trial was subcutaneous, while it was intravenously administered in the CORIMUNO-ANA-1 trial. Both did not show any significant benefits in their defined outcomes. Whether higher doses or extended duration of anakinra could have benefited the COVID-19 patients is unknown and may warrant further investigation.

This trial has some limitations. First, a placebo was not used in this trial due to the nature of the pandemic and the urgency to answer the clinical question. In addition, there is a potential for bias due to the open-label nature of the trial. However, this was minimized by choosing objective outcomes and blinding the outcome assessors. Finally, the overall sample size was small, especially for patients with severe disease or mechanical ventilation; hence, subgroups analysis in this population was not feasible.

On the other hand, this trial has several strengths. This trial is one of the few randomized trials conducted on COVID-19 patients after establishing a better SOC (steroids, remdesivir). Moreover, it was a multicenter trial involving multiple ethnic groups, contributing to the study results’ higher generalizability. The WHO’s recommended outcomes for clinical studies were used; those included the WHO clinical progression score, viral burden as measured by the cyclic threshold, and the overall mortality, which facilitate the comparison of this trial’s results to other similar trials. Furthermore, this trial was conducted in Qatar where well-established national guidelines for COVID-19 treatment exist. Finally, no significant loss of follow-up rate was reported in this study.

## Conclusion

In patients with severe COVID-19 pneumonia requiring oxygen therapy, the addition of anakinra to the SOC treatment was not associated with significant improvement in the WHO clinical progression scale. Therefore, there is a need for determining the subgroups of COVID-19 patients who will benefit the most by the timely administration of anakinra.

## Data Availability

All relevant data are within the manuscript and its Supporting Information files.

## Author Contributions

Conceptualization and Methodology: *Eman Elmekaty*. Data Validation: *Mohamed Aboukamar, Waqar Munir, Rim Alibrahim, Eman Elmekaty. Formal analysis and Visualization: Mohamed Izham*. Investigations: *Waqar Munir, Ahmed Al Bishawi, Arun Nair, Aya Maklad, Rawan Abouelhassan, Fatima Iqbal, Mohammed AbuKhattab, Rim Alibrahim, Mohamed Aboukamar, Alaaeldin Abdelmajid*. Resources: *Hussam Al Soub, Mohammed AbuKhattab, Muna Al Maslamani*. Data curation: *Aya Maklad, Rawan Abouelhassan, Eman Elmekaty*. Writing – original draft: *Ahmed Al Bishawi, Arun Nair, Mohamed Izham, Fatima Iqbal*. Writing – review & editing: *Aya Maklad, Rawan Abouelhassan, Mohamed AbuKhattab, Eman Elmekaty, Hussam Al Soub, Alaaeldin Abdelmajid*. Supervision: *Hussam Al Soub, Muna Al Maslamani*.

## Data sharing

Deidentified participants data (in spreadsheet form, with included data definitions) will be made available for sharing after publication on reasonable requests made to the corresponding author. Data can be shared through secure online platforms after being approved by the principal investigator.

## Acknowledgments

We would like to thank all physician assistants working in Hamad Medical Corporation COVID-19 facilities for their outstanding support and dedication.

## References

1. Lai CC, Shih TP, Ko WC, Tang HJ, Hsueh PR. Severe acute respiratory syndrome coronavirus 2 (SARS-CoV-2) and coronavirus disease-2019 (COVID-19): The epidemic and the challenges. Int J Antimicrob Agents. 2020 Feb 17:105924. doi: 10.1016/j.ijantimicag.2020.105924. [Epub ahead of print]

2. WHO Coronavirus Disease (COVID-19) Dashboard. World Health Organization, 2020. https://Covid19.who.int/. [Accessed 29 March 2020.]

3. Guan W-j, Ni Z-y, Hu Y, et al. Clinical characteristics of 2019 novel coronavirus infection in China. MedRxiv 2020: 2020.02.06.20020974.

4. WHO Director-General’s opening remarks at the media briefing on COVID-19 - 3 March 2020 - World Health Organization, March 3, 2020

5. Chen G, Wu D, Guo W, et al. Clinical and immunological features of severe and moderate coronavirus disease 2019. J Clin Invest 2020;130:2620–9. doi:10.1172/JCI137244

6. Mehta P, McAuley DF, Brown M, et al. COVID-19: consider cytokine storm syndromes and immunosuppression. Lancet 2020; 395: 1033–34.

7. McGonagle D, Sharif K, O’Regan A, Bridgewood C. The role of cytokines including interleukin-6 in COVID-19 induced pneumonia and macrophage activation syndrome-like disease. Autoimmun Rev 2020; published online April 3. DOI:10.1016/j.autrev.2020.102537.

8. Shakoory B, Carcillo JA, Chatham WW, Amdur RL, Zhao H, Dinarello CA, et al. Interleukin-1 receptor blockade is associated with reduced mortality in sepsis patients with features of macrophage activation syndrome: reanalysis of a prior phase III trial. Crit Care Med. 2016;44:275– 81.

9. Opal SM, Fisher Jr CJ, Dhainaut JF, Vincent JL, Brase R, Lowry SF, et al. Confirmatory interleukin-1 receptor antagonist trial in severe sepsis: a phase III, randomized, double-blind, placebo-controlled, multicenter trial. Crit Care Med. 1997;25:1115–24.

10. Huet T, Beaussier H, Voisin O, et al. Anakinra for severe forms of COVID-19: a cohort study. Lancet Rheumatol. 2020;9913(20):1–8. doi:10.1016/s2665-9913(20)30164-8

11. Cavalli G, De Luca G, Campochiaro C, et al. Interleukin-1 blockade with high-dose anakinra in patients with COVID-19, acute respiratory distress syndrome, and hyperinflammation: a retrospective cohort study. Lancet Rheumatol. 2020;2(6):e325–e331. doi:10.1016/S2665-9913(20)30127-2

12. Langer-Gould A, Smith JB, Gonzales EG, et al. Early identification of COVID-19 cytokine storm and treatment with anakinra or tocilizumab. Int J Infect Dis. 2020;99:291–297. doi:10.1016/j.ijid.2020.07.08

13. Marshall JC, Murthy S, Diaz J, Adhikari NK, Angus DC, Arabi YM, et al. A minimal common outcome measure set for COVID-19 clinical research. The Lancet Infectious Diseases. 2020;20(8):e192–e7.

14. National Cancer Institute. Common Terminology Criteria for Adverse Events - Version 5.0. 2017. Available from: https://ctep.cancer.gov/protocoldevelopment/electronic_applications/docs/ctcae_v5_quick_reference_5x7.pdf

15. Effect of anakinra versus usual care in adults in hospital with COVID-19 and mild-to-moderate pneumonia (CORIMUNO-ANA-1): a randomized controlled trial. Lancet Respir Med. 2021 Mar;9(3):295–304.

16. Rajasekaran S, Kruse K, Kovey K, Davis AT, Hassan NE, Ndika AN, et al. Therapeutic role of anakinra, an interleukin-1 receptor antagonist, in the management of secondary hemophagocytic lymphohistiocytosis/sepsis/multiple organ dysfunction/macrophage activating syndrome in critically ill children*. Pediatr Crit Care Med J Soc Crit Care Med World Fed Pediatr Intensive Crit Care Soc. 2014 Jun;15(5):401–8.

17. Mehta P, Cron RQ, Hartwell J, Manson JJ, Tattersall RS. Silencing the cytokine storm: the use of intravenous anakinra in haemophagocytic lymphohistiocytosis or macrophage activation syndrome. Lancet Rheumatol. 2020 Jun;2(6):e358–67.

18. Zeng F, Huang Y, Guo Y, Yin M, Chen X, Xiao L, et al. Association of inflammatory markers with the severity of COVID-19: A meta-analysis. Int J Infect Dis. 2020 Jul;96:467–74.

19. Navarro-Millán I, Sattui SE, Lakhanpal A, Zisa D, Siegel CH, Crow MK. Use of Anakinra to Prevent Mechanical Ventilation in Severe COVID-19: A Case Series. Arthritis Rheumatol Hoboken Nj. 2020 Jun 30;10.1002/art.41422.

20. Aouba A, Baldolli A, Geffray L, Verdon R, Bergot E, Martin-Silva N, et al. Targeting the inflammatory cascade with anakinra in moderate to severe COVID-19 pneumonia: case series. Ann Rheum Dis. 2020 Oct;79(10):1381–2.

21. Dimopoulos G, de Mast Q, Markou N, Theodorakopoulou M, Komnos A, Mouktaroudi M, et al. Favorable Anakinra Responses in Severe Covid-19 Patients with Secondary Hemophagocytic Lymphohistiocytosis. Cell Host Microbe. 2020 Jul 8;28(1):117–123.e1.

22. The Safety and Efficacy of Anakinra, an Interleukin-1 Antagonist in Severe Cases of COVID-19: A Systematic Review and Meta-Analysis [Internet]. [cited 2021 Aug 14]. Available from: https://www.ncbi.nlm.nih.gov/pmc/articles/PMC8258297/

23. The RECOVERY Collaborative Group. Dexamethasone in Hospitalized Patients with Covid-19 — Preliminary Report. N Engl J Med. 2020 Jul 17;NEJMoa2021436.

24. Bhimraj A, Morgan RL, Shumaker AH, Lavergne V, Baden L, Cheng VC-C, et al. Infectious Diseases Society of America Guidelines on the Treatment and Management of Patients with COVID-19. Clin Infect Dis Off Publ Infect Dis Soc Am. 2020 Apr 27;ciaa478.

25. Rao SN, Manissero D, Steele VR, Pareja J. A Narrative Systematic Review of the Clinical Utility of Cycle Threshold Values in the Context of COVID-19. Infect Dis Ther. 2020 Sep;9(3):573–86.

26. Cavalli G, Dagna L. The right place for IL-1 inhibition in COVID-19. Lancet Respir Med. 2021 Mar;9(3):223–4.

27. Hellenic Institute for the Study of Sepsis. suPAR-Guided Anakinra Treatment for Validation of the Risk and Early Management of Severe Respiratory Failure by COVID-19: The SAVE-MORE Double-blind, Randomized, Phase III Confirmatory Trial [Internet]. https://clinicaltrials.gov; 2021 Apr [cited 2021 Aug 12]. Report No.: NCT04680949. Available from: https://clinicaltrials.gov/ct2/show/NCT04680949

28. Rovina N, Akinosoglou K, Eugen-Olsen J, Hayek S, Reiser J, Giamarellos-Bourboulis EJ. Soluble urokinase plasminogen activator receptor (suPAR) as an early predictor of severe respiratory failure in patients with COVID-19 pneumonia. Crit Care. 2020 Apr 30;24:187.

29. Kyriazopoulou E, Panagopoulos P, Metallidis S, Dalekos GN, Poulakou G, Gatselis N, et al. An open label trial of anakinra to prevent respiratory failure in COVID-19. eLife. 10:e66125.

30. Dalekos GN, Stefos A, Georgiadou S, Lygoura V, Michail A, Ntaios G, et al. Lessons from pathophysiology: Use of individualized combination treatments with immune interventional agents to tackle severe respiratory failure in patients with COVID-19. Eur J Intern Med. 2021 Jun;88:52–62.

